# Vascular risk factors mediate the relationship between education and white matter hyperintensities

**DOI:** 10.1101/2025.06.03.25328899

**Authors:** Shima Raeesi, Yashar Zeighami, Roqaie Moqadam, Cassandra Morrison, Mahsa Dadar

## Abstract

**INTRODUCTION:** Education can protect against cognitive decline and dementia through cognitive reserve and reduced vascular risk. This study examined whether vascular risk mediates the relationship between education and white matter hyperintensity (WMH) burden.

**METHODS:** Data from 1089 older adults from the National Alzheimer’s Coordinating Center were analyzed. A composite vascular score was created using diabetes, hypertension, hypercholesterolemia, smoking, alcohol abuse, body mass index, and blood pressure. Linear regressions and mediation analyses examined associations and indirect effects between education, vascular risk, and WMH, adjusting for age, sex, and diagnosis.

**RESULTS:** Higher education was associated with lower vascular risk (*p* < .001) and WMH burden (*p* = .01). Mediation analysis showed an indirect effect of education on WMH via vascular risk (a*b = −0.02, *p* = .004), accounting for 23% of the total effect.

**DISCUSSION:** Education influences cerebrovascular health via reducing vascular risk. Addressing vascular health may reduce WMH burden.

## 1. Background

White matter hyperintensities (WMHs) are magnetic resonance imaging (MRI) detectable lesions indicative of cerebrovascular disease (1). Their presence is strongly associated with cognitive decline and an increased risk of dementia, including Alzheimer’s disease (AD) (2–6). WMHs often present as hyperintense regions on T2-weighted and Fluid-attenuated inversion recovery (FLAIR) MRI scans and are thought to reflect chronic ischemia and microvascular damage (3,7). WMHs are particularly common among older adults, increasing in prevalence with age (3,8–10), and becoming more extensive in individuals with vascular comorbidities, such as hypertension and diabetes (11–14).

WMHs are increasingly recognized as a biomarker of cerebral small vessel disease, however, the pathophysiology of WMHs is complex and multifactorial, involving chronic hypoperfusion, endothelial dysfunction, and small vessel pathology (7,15). Evidence from neuropathological studies indicates that WMHs may arise from a combination of ischemic demyelination, gliosis, and axonal loss (16,17). Some studies suggest that WMHs may represent a continuum of cerebrovascular injury, ranging from mild, asymptomatic changes to extensive, clinically significant damage. WMH burden is also predictive of future cognitive decline, progression to mild cognitive impairment (MCI), and an increased risk of dementia (9,18,19). Finally, the spatial distribution of WMHs have been shown to relate differently to dementia and vascular risk factors. For example, frontal WMHs are more closely linked to vascular risk factors whereas parietal is more strongly linked to dementia risk and conversion (16,20,21).

Education has long been recognized as a protective factor against cognitive decline, a concept often attributed to the cognitive reserve hypothesis (22–25). This hypothesis posits that individuals with higher educational attainment develop more robust cognitive processes or neural networks, allowing them to better cope with age-related brain changes and pathology, compared to people with lower education (25–27). However, the mechanisms through which education confers this protective effect are multifaceted and not entirely understood. While some studies emphasize the role of lifelong cognitive engagement, others highlight the potential for education to modify health behaviors, thereby reducing vascular risk (26,28,29). Emerging evidence suggests that education may influence health behaviors and lifestyle choices, leading to reduced exposure to vascular risk factors (29–31). For instance, individuals with higher education levels are more likely to engage in health-promoting behaviors, have better access to healthcare resources, and possess greater health literacy, all of which contribute to lower incidences of different vascular conditions (29,30,32). These factors may collectively reduce the burden of WMHs and subsequent cognitive decline (21). High educational attainment is linked to better cardiovascular health, both of which are critical in reducing cerebrovascular damage. Individuals with higher education demonstrate slower WMH progression over time, even when controlling for age and vascular risk factors. (33).

There is increasing interest in the potential mediation effect of vascular risk factors on the relationship between education and WMH burden. Understanding the interplay between education, vascular risk, and WMHs is crucial, as it underscores the potential of educational interventions and public health strategies aimed at reducing vascular risks to mitigate WMH accumulation and preserve cognitive function (21, 22). Given the increasing prevalence of dementia and cerebrovascular diseases, identifying modifiable factors that influence WMH progression remains a public health priority (23). While education appears to offer a protective effect against cognitive decline, this benefit is likely influenced by a complex interplay of factors, including vascular health. Investigating the mediating role of vascular risk factors in the relationship between education and WMH burden can provide valuable insights into preventive strategies for cognitive impairment and dementia (24).

The primary objective of this study was to investigate whether vascular risk factors mediate the relationship between education and WMH volume. Data were drawn from the National Alzheimer’s Coordinating Center (NACC, https://naccdata.org/) dataset (34), which includes comprehensive clinical, MRI, and neuropsychological assessments from multiple sites across the United States, providing a robust, community-based representation of aging and cognitive health.

## 2. Methods

### 2.1 National Alzheimer’s Coordinating Center Participants

Data were obtained from the NACC database, including the NACC Uniform Data Set (UDS) and MRI Data Set, with T1-weighted and FLAIR MRI scans available to extract regional WMH measures (34–36). Baseline data from individuals aged 55 and over with diagnoses of cognitively normal, amnestic MCI, non-amnestic MCI, and AD were included in the study. Cognitive diagnoses were derived using the NACCTMCI and NACCETPR variables, which represent the primary diagnosis. Participants were selected based on the availability of MRI data and vascular risk factor information. Education was quantified as years of formal schooling as a numeric variable. Due to significant differences in education levels across racial groups, race-stratified analyses were considered. However, because of limited sample sizes and reduced statistical power in the non-White subgroups, the main analyses were conducted within the White participant group (n = 1,089), which provided sufficient data for robust modeling. Exploratory results from the non-White groups are presented separately in the Supplementary Materials.

### 2.2 MRI and Volumetric WMH Measurements

MRI Preprocessing: T1-weighted and FLAIR MRI scans were processed using an open-source image processing pipeline, integrating tools from the MINC Toolkit v2 (MINC Toolkit) and Advanced Normalization Tools (ANTs) Toolkit (37). The preprocessing pipeline included essential steps such as noise reduction (38), correction for intensity non-uniformity (39), and intensity normalization to a standardized range of [0–100]. After initial preprocessing, the T1-weighted images underwent linear registration (40) to the MNI-ICBM152 average template (41) using nine parameters (translation, rotation, and scaling) to correct for head size and orientation differences, followed by a nonlinear registration (42). WMH volumes were identified based on T1-weighted and FLAIR images using BISON (43), an automated segmentation tool previously validated for WMH detection in other multi-center studies as well as in NACC (44–47). This method leverages a Random Forest classifier combined with both location-specific features and intensity features, which are derived from a comprehensive library of manually segmented scans. All processing steps, including linear and nonlinear registration and WMH segmentation were visually quality controlled by an experienced rater (RM), blind to clinical diagnosis. Cases that failed to meet quality control criteria (N = 28) were omitted from further analysis. WMH volume was quantified as the number of voxels labeled as WMH within the standard space (mm³), i.e. normalized for intracranial volume. Regional WMH volumes were calculated separately for each brain lobe (frontal, temporal, parietal, and occipital) and hemisphere using the Hammer’s lobar atlas (48,49). Figure 1 shows an example of the WMH segmentations for 3 NACC participants. To achieve a normal distribution, the calculated WMH volumes were log-transformed prior to statistical analysis.

**Figure 1.**
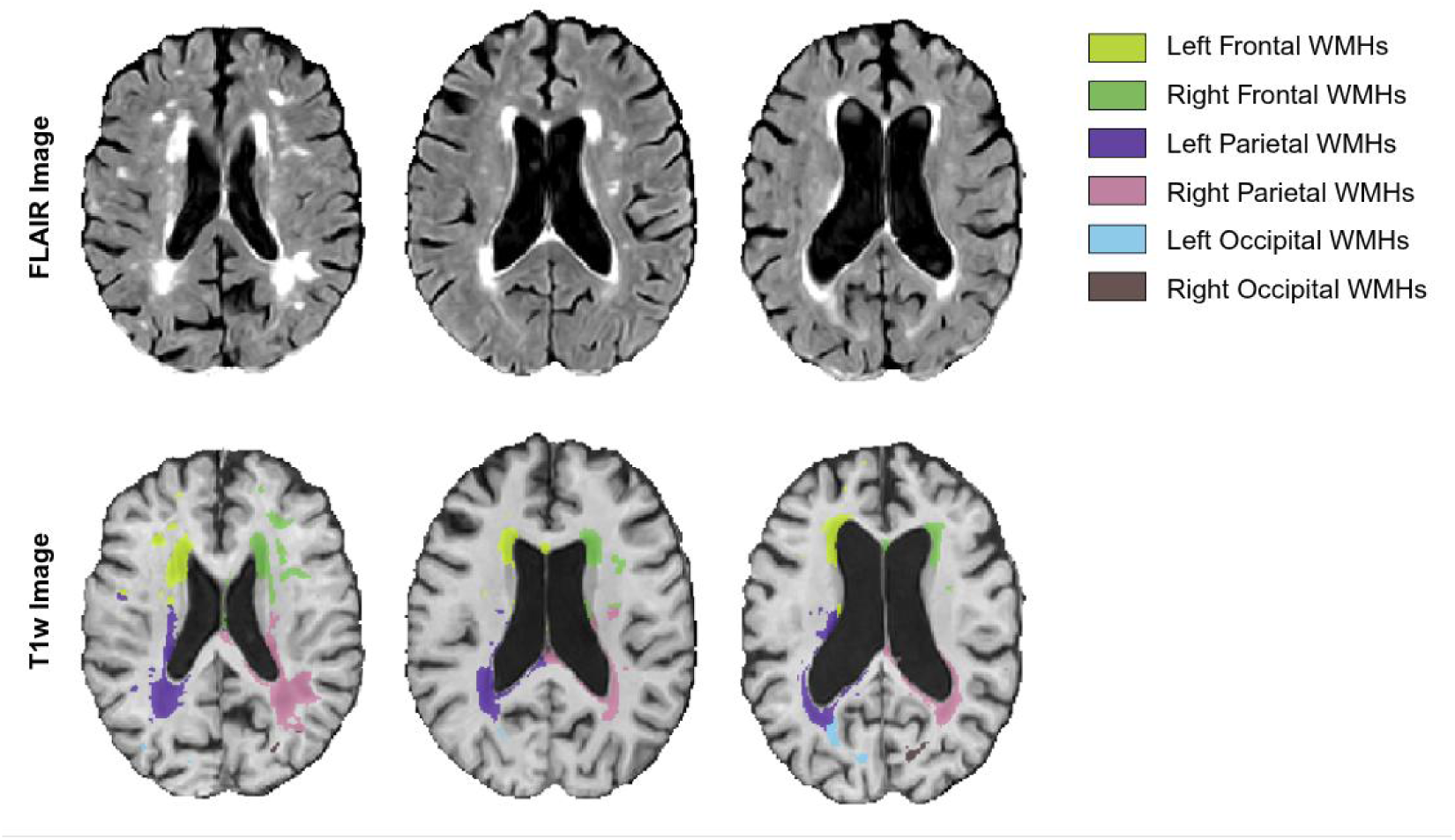
Illustrative examples of WMH segmentations derived from FLAIR and T1-weighted MRI scans across three participants with varying WMH burden. WMH = White Matter Hyperintensity. FLAIR = Fluid-Attenuated Inversion Recovery MRI scans.

### 2.3 Vascular Risk Factors

To comprehensively evaluate the influence of vascular risk factors on WMH burden, we systematically assessed multiple variables reflecting vascular health. Diabetes status was determined using values from the DIABET and DIABETES variables based on health history. Hypertension was identified using the HYPERT and HYPERTEN variables. Hypercholesterolemia was derived from the HYPERCHO and HYPERCHOL variables. Alcohol abuse was assessed using the ALCOHOL variable, which was defined as alcohol abuse causing clinically significant impairment in areas such as work, driving, legal, or social domains over a 12-month period. For all these variables, binary coding was used, where ‘0’ indicated the absence and ‘1’ indicated the presence of the risk factor.

In contrast, the following vascular indicators were treated as continuous variables: Body Mass Index (BMI), calculated using the NACCBMI column based on recorded height and weight; systolic and diastolic blood pressure, derived from the BPSYS and BPDIAS columns, respectively; and smoking behavior, which was quantified using total years smoked (SMOKYRS) and average number of packs smoked per day (PACKSPER). These continuous variables allowed for more granular assessment of vascular risk burden and were analyzed accordingly.

### 2.4 Vascular Composite Score

To facilitate mediation analysis and assess the cumulative impact of vascular risk factors, we developed a vascular composite score (50–53). This score allowed for the evaluation of whether the collective vascular risk mediated the relationship between education and WMHs. To achieve uniform integration within the composite score, continuous variables such as BMI, systolic blood pressure, and diastolic blood pressure were transformed into binary variables. Following the guidelines from the National Institute of Health and the National Institute on Aging for older adults, as well as the 2017 ACC/AHA Guideline for the Prevention, Detection, Evaluation, and Management of High Blood Pressure in Adults, high systolic blood pressure was defined as ≥130 mmHg and high diastolic blood pressure as ≥80 mmHg (54,55). Based on the Centers for Disease Control and Prevention, a BMI of 25 or more is categorized as high (overweight or obesity) (28). Consequently, for these three variables, a value of ‘1’ indicates elevated levels, while ‘0’ indicates normal levels. Smoking status was also incorporated into the composite score, in which individuals with any history of smoking were coded as ‘1’, and non-smokers were coded as ‘0’. The vascular composite score was then calculated by summing the presence (1) or absence (0) of the eight binary-coded vascular risk factors (diabetes, hypertension, hypercholesterolemia, smoking history, alcohol abuse, high BMI, high systolic, and high diastolic blood pressure), resulting in a continuous measure ranging from 0 to 8 per participant. This composite score facilitated comprehensive mediation analysis within the study framework, allowing for a more integrated assessment of the mediating role of vascular risk on the relationship between education and WMH burden.

### 2.5 Statistical Analysis

Demographic and clinical characteristics were analyzed using independent sample t-tests and chi-square tests, with false discovery rate (FDR) correction applied for multiple comparisons. All analyses were conducted in MATLAB R2024b. To investigate the relationship between education and individual vascular risk factors, we employed a series of linear regression models, adjusting for covariates including age, sex, and diagnostic status. Vascular risk factors were modeled based on their measurement level: categorical variables (e.g., presence or absence of diabetes, hypertension, hypercholesterolemia, alcohol abuse) were treated as binary outcomes, while continuous variables (e.g., BMI, systolic and diastolic blood pressure, smoking years, and packs per day) were treated as continuous outcomes. Accordingly, linear regression was used for continuous outcomes, and logistic regression was applied to binary outcomes to ensure appropriate modeling for each variable type.

In both approaches, continuous predictors were z-scored prior to analysis to standardize their distributions and facilitate interpretability. Each vascular risk factor was analyzed independently to estimate its association with education. The model equation for continuous outcomes can be expressed as:

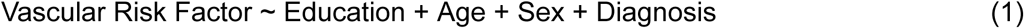

Effect sizes (estimates) were computed to assess whether the observed associations differed significantly from zero. Statistical significance was defined as FDR adjusted (56) *p* < 0.05 to account for multiple comparisons. All p-values reported in the text reflect values after FDR correction.

Additional analyses were conducted to evaluate the relationship between education and WMH volume. WMH volumes were log-transformed to achieve a more normal distribution and z-scored prior to the analyses. This analysis was completed for each region of interest including total, and right and left frontal, temporal, parietal, and occipital. Thus, the primary dependent variable was the log-transformed WMH volume, while the independent variable of interest was education.

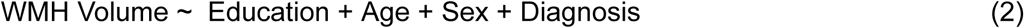

To investigate the mediating effect of vascular risk factors on the relationship between education and WMH burden, a mediation model was estimated using the Structural Equation Modeling (SEM) approach in the *mediation* package in R (57). All variables were standardized (z-scored) before inclusion. Age, sex, and diagnosis were included as covariates. To assess the significance of the indirect effects, bootstrapping was performed with 1,000 resamples The proportion of the total effect mediated was calculated to assess the relative contribution of the indirect pathway compared to the direct effect.

## 3 Results

### 3.1 Demographics and Clinical Data

Table 1 summarizes the demographic, clinical, vascular, and neuroimaging characteristics of the study participants. The sample consisted of 1,089 White older adults, including 505 cognitively normal (NC) individuals, 319 with MCI, and 265 with AD. The mean age of the participants was 73.86 ± 8.49. NC participants were significantly younger (71.91 ± 8.26) than those with MCI (75.94 ± 8.04, *p* < .001) and AD (75.05 ± 8.66, *p* < .001). The mean education level was 15.87 ± 2.86, with the highest educational attainment observed in the NC group (16.19 ± 2.62), which was significantly greater than both MCI (*p* < .004) and AD (*p* < .016). Education levels did not differ significantly (*p* = .69) between MCI (15.55 ± 3.12) and AD (15.65 ± 3.07). The cohort was 57% female, with a significantly higher proportion of females in the NC group compared to both MCI and AD groups (*p* < .001); no significant sex difference was observed between MCI and AD.

**Table 1:**
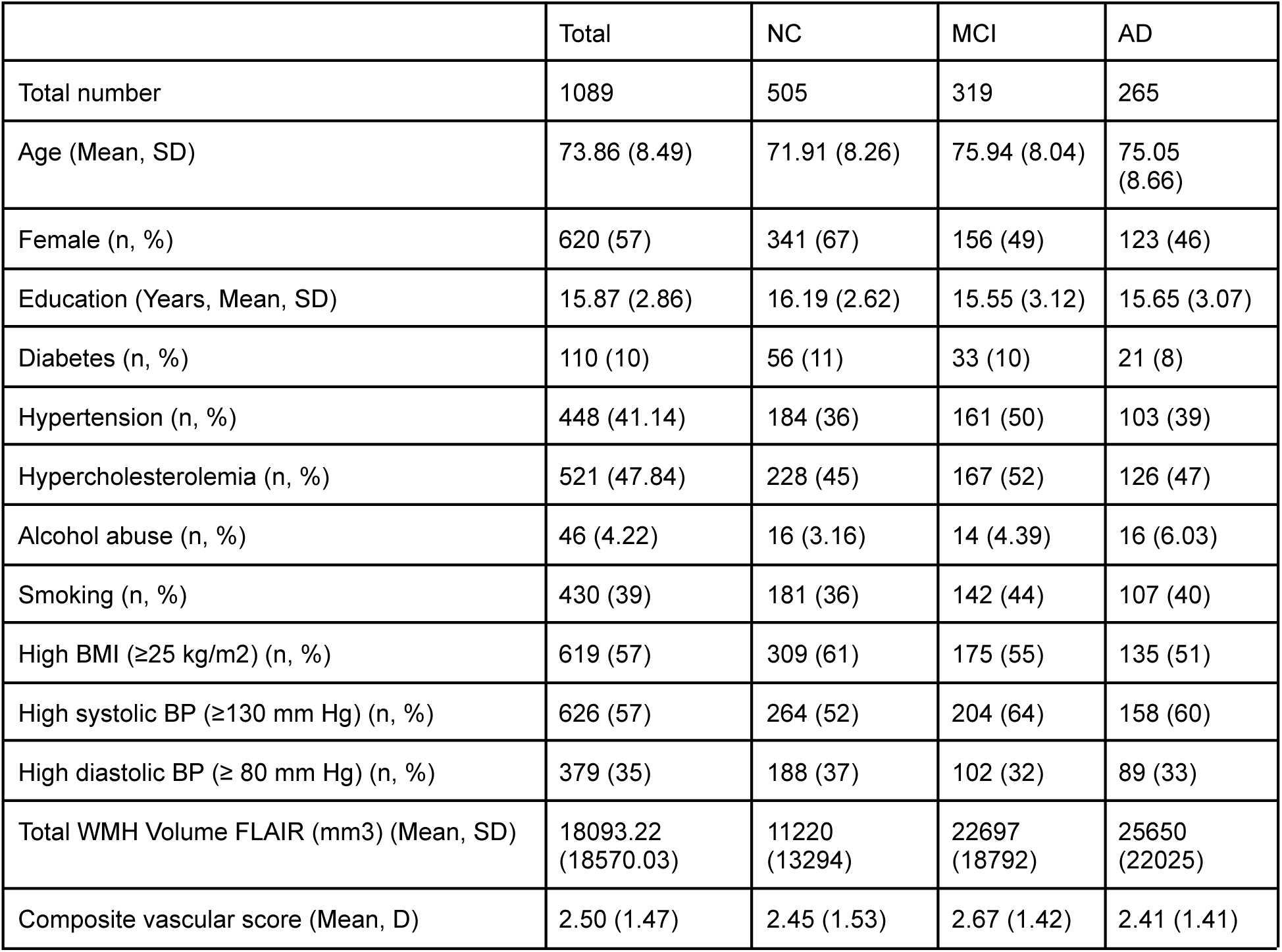
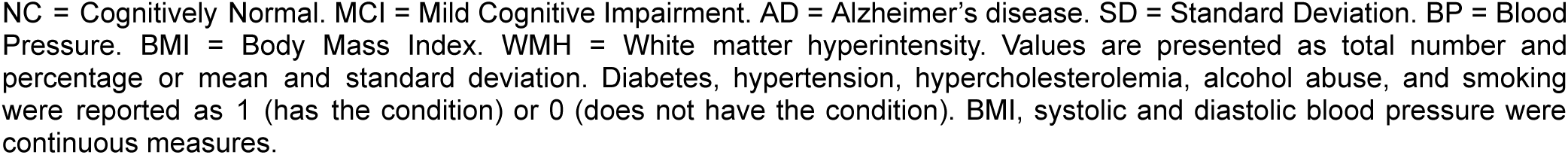
Demographic, Clinical, and Neuroimaging Characteristics for Study Participants.

|  | Total | NC | MCI | AD |
| --- | --- | --- | --- | --- |
| Total number | 1089 | 505 | 319 | 265 |
| Age (Mean, SD) | 73.86 (8.49) | 71.91 (8.26) | 75.94 (8.04) | 75.05 (8.66) |
| Female (n, %) | 620 (57) | 341 (67) | 156 (49) | 123 (46) |
| Education (Years, Mean, SD) | 15.87 (2.86) | 16.19 (2.62) | 15.55 (3.12) | 15.65 (3.07) |
| Diabetes (n, %) | 110 (10) | 56 (11) | 33 (10) | 21 (8) |
| Hypertension (n, %) | 448 (41.14) | 184 (36) | 161 (50) | 103 (39) |
| Hypercholesterolemia (n, %) | 521 (47.84) | 228 (45) | 167 (52) | 126 (47) |
| Alcohol abuse (n, %) | 46 (4.22) | 16 (3.16) | 14 (4.39) | 16 (6.03) |
| Smoking (n, %) | 430 (39) | 181 (36) | 142 (44) | 107 (40) |
| High BMI ($\geq 25$ kg/m<sup>2</sup>) (n, %) | 619 (57) | 309 (61) | 175 (55) | 135 (51) |
| High systolic BP ($\geq 130$ mm Hg) (n, %) | 626 (57) | 264 (52) | 204 (64) | 158 (60) |
| High diastolic BP ($\geq 80$ mm Hg) (n, %) | 379 (35) | 188 (37) | 102 (32) | 89 (33) |
| Total WMH Volume FLAIR (mm<sup>3</sup>) (Mean, SD) | 18093.22 (18570.03) | 11220 (13294) | 22697 (18792) | 25650 (22025) |
| Composite vascular score (Mean, D) | 2.50 (1.47) | 2.45 (1.53) | 2.67 (1.42) | 2.41 (1.41) |

NC = Cognitively Normal. MCI = Mild Cognitive Impairment. AD = Alzheimer's disease. SD = Standard Deviation. BP = Blood Pressure. BMI = Body Mass Index. WMH = White matter hyperintensity. Values are presented as total number and percentage or mean and standard deviation. Diabetes, hypertension, hypercholesterolemia, alcohol abuse, and smoking were reported as 1 (has the condition) or 0 (does not have the condition). BMI, systolic and diastolic blood pressure were continuous measures.

Among vascular risk factors, hypertension (41.14%) and hypercholesterolemia (47.84%) were highly prevalent. There were no significant differences in the prevalence of these vascular risk factors across diagnostic groups. More than half of the participants (57%) had BMI ≥25 kg/m², with the highest proportion observed in the NC group (61%), and group means as follows: NC (27.36 ± 5.15), MCI (26.49 ± 4.78), and AD (26.03 ± 4.45). BMI was significantly lower in the MCI group compared to NC (p < .02), and significantly lower in the AD group compared to NC (*p* < .001). High systolic blood pressure (≥130 mm Hg) was present in 57%, while high diastolic blood pressure (≥80 mm Hg) was observed in 35%, with no significant differences between groups. In terms of neuroimaging findings, the mean total WMH volume was 18.09 CCs (SD = 18.57), with the highest values observed in the AD group (25.65 CCs) compared to NC (11.22 CCs, *p* < .001). There were no significant differences in WMH volumes between MCI and AD participants (*p* = .08). The mean composite vascular score across the cohort was 2.50 (SD = 1.47), reflecting a moderate burden of cumulative vascular risk, with no significant group differences. Characteristics of the non-white participants are presented in Tables S1-3 in the supplementary materials.

### 3.2 Education and Vascular Risk Factors

Higher educational attainment was significantly associated with lower systolic blood pressure (β = −0.09, *p* = .003), reduced BMI (β = −0.14, *p* < .001), and decreased smoking exposure (β = −0.15, *p* < .001. Additionally, individuals with more years of education had lower odds of diabetes (β = −0.12, *p* = 0.007), hypertension (β = −0.15, *p* = .003), and hypercholesterolemia (β = −0.16, *p* = .002). No significant associations were observed for diastolic blood pressure or alcohol abuse. There was also a significant inverse relationship between education and the composite vascular risk score (β = –0.09, *p* < .001. Detailed results are presented in Table 2. The associations between education and vascular risk factors for the non-white participants are presented in Supplementary Table 4.

**Table 2:** Associations Between Educational Attainment and Individual Vascular Risk Factors.

|  | Estimate | tStat | *p* Value |
| --- | --- | --- | --- |
| Diabetes | - 0.12 | - 2.68 | **.007** |
| Hypertension | - 0.15 | - 2.92 | **.003** |
| Systolic Blood Pressure | - 0.09 | - 2.94 | **.003** |
| Diastolic Blood Pressure | 0.03 | - 1.03 | .29 |
| Hypercholesterolemia | - 0.16 | - 3.02 | **.002** |
| Alcohol Abuse | - 0.11 | - 2.66 | .07 |
| Smoking (Years) | - 0.15 | - 4.77 | **< .001** |
| Smoking (Packs Per Day) | - 0.16 | - 4.90 | **< .001** |
| BMI | - 0.14 | - 4.68 | **< .001** |

BMI = Body Mass Index. *Bolded values are those that remain significant after correction for multiple comparisons.*

### 3.3 Education and Regional WMH Burden

Linear regression analysis revealed that higher educational attainment was significantly associated with lower total WMH burden (β = −0.09, *p* = .01) after adjusting for key covariates, including age, sex and cognitive status, indicating that education contributes independently to cerebrovascular health. Table 3 presents the results of region-specific linear regression models evaluating the association between education and WMH burden across bilateral frontal, temporal, parietal, and occipital lobes. Significant negative associations between education and WMH volume were observed in total WMH, right frontal, left and right temporal and parietal lobes (β belongs to [−.08, −.12], *p =* .01). In contrast, occipital WMH volumes were not significantly associated with education in either hemisphere (*p* > 0.1). Associations between education and total as well as regional WMHs were not statistically significant in non-white participants (Table 5 in the supplementary materials).

**Table 3:** Associations Between Education and Regional WMH Volumes.

|  | Estimate | tStat | p value |
| --- | --- | --- | --- |
| Total WMH Volume | -0.09 | -2.66 | **0.01** |
| L frontal WMH Volume | -0.07 | -1.94 | 0.05 |
| R frontal WMH Volume | -0.08 | -2.60 | **0.01** |
| L temporal WMH Volume | -0.12 | -2.59 | **0.01** |
| R temporal WMH Volume | -0.12 | -2.66 | **0.01** |
| L Occipital WMH Volume | -0.07 | -1.41 | 0.16 |
| R Occipital WMH Volume | -0.04 | -0.81 | 0.42 |
| L parietal WMH Volume | -0.11 | -2.54 | **0.01** |
| R parietal WMH Volume | -0.11 | -2.63 | **0.01** |

*L= Left Hemisphere. R = Right Hemisphere. Bolded values are those that remained significant after correction for multiple comparisons.*

### 3.4 Mediation Analysis of Education, Vascular Risk, and Regional WMH Burden

Mediation analysis was conducted to examine whether vascular risk factors mediate the relationship between educational attainment and total as well as regional WMH burden. The models assessed both the direct effect of education on WMH volume and the indirect effect via a composite vascular risk score. For total WMHs, there was evidence of full mediation, with a non-significant direct effect of education on WMH burden (c’ = −0.05, *p* = .114) and a significant indirect effect through vascular risk factors (a*b = −0.02, *p* = .004), accounting for 23% of the total effect (c = −0.065, *p* = .044). Figure 2 illustrates this mediation model, showing the significant pathways from education to vascular risk and from vascular risk to WMH burden. Table 4 presents the results of region-specific mediation analyses examining whether vascular risk factors mediate the relationship between educational attainment and regional WMH burden. In bilateral frontal and left temporal regions, indirect effects were significant (β belongs to [−.02, −.11], *p <* .006), while the direct effects were non-significant, indicating that the association between education and WMH burden in these regions is primarily mediated by vascular risk factors (i.e. a full mediation). The composite score was most strongly associated with WMH burden in the left frontal lobe (β = 0.10, *p* = <.001), supporting the hypothesis that vascular burden significantly contributes to WMH accumulation in this region. The proportion of the total effect mediated ranged from 21% to 41%, with the highest observed in the left frontal region. These findings suggest that vascular health plays a key role in linking education to WMH accumulation specifically in anterior regions, supporting the hypothesis of a modifiable vascular pathway in these areas.

**Figure 2:**
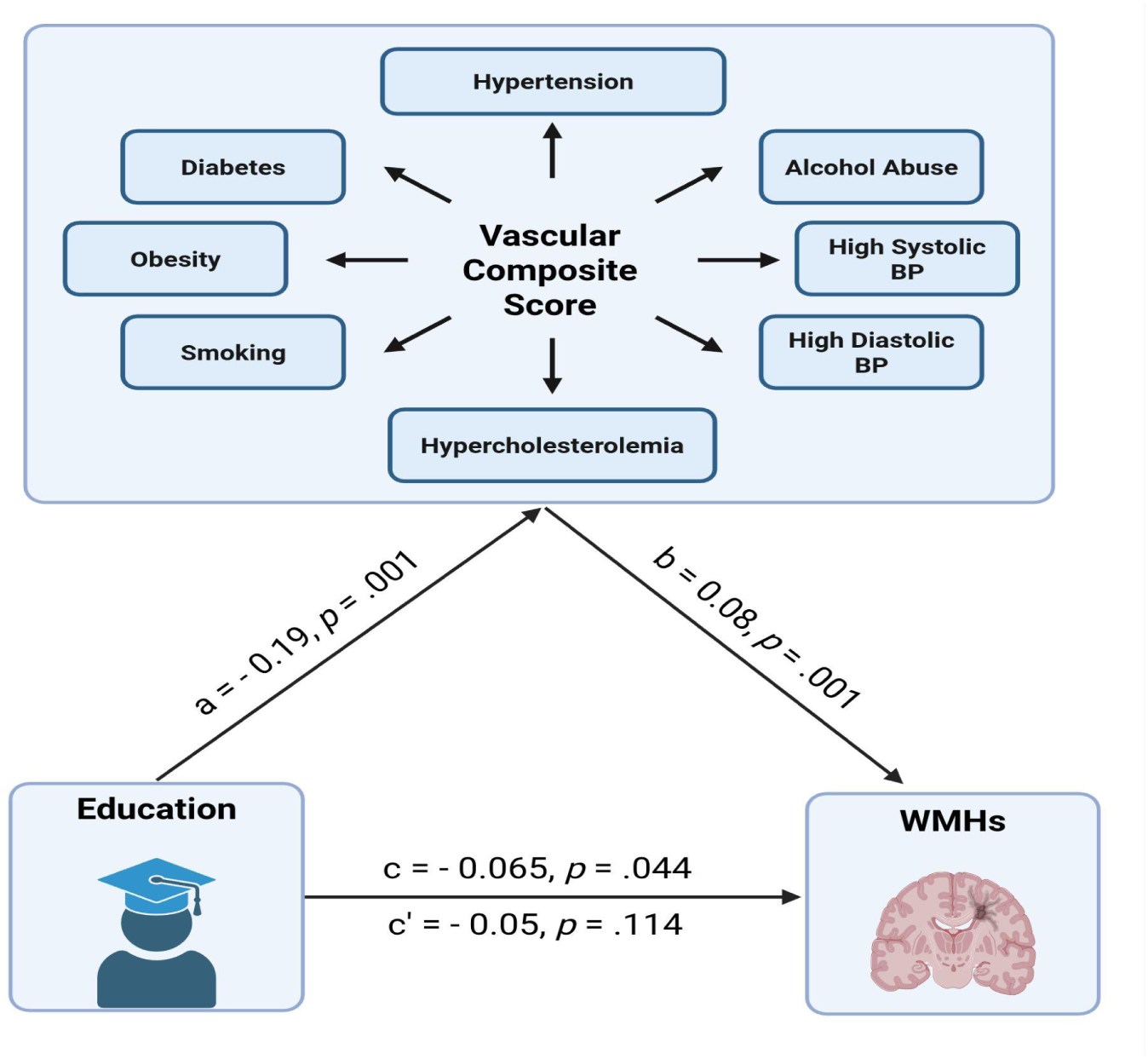
Mediation model of the relationship between education and WMH burden via composite vascular risk score. WMH = White Matter Hyperintensity, BP = Blood Pressure. Mediation model showing that vascular risk partially mediates the relationship between education and WMH burden. All pathways were significant, with approximately 22% of the total effect mediated.

**Table 4:**
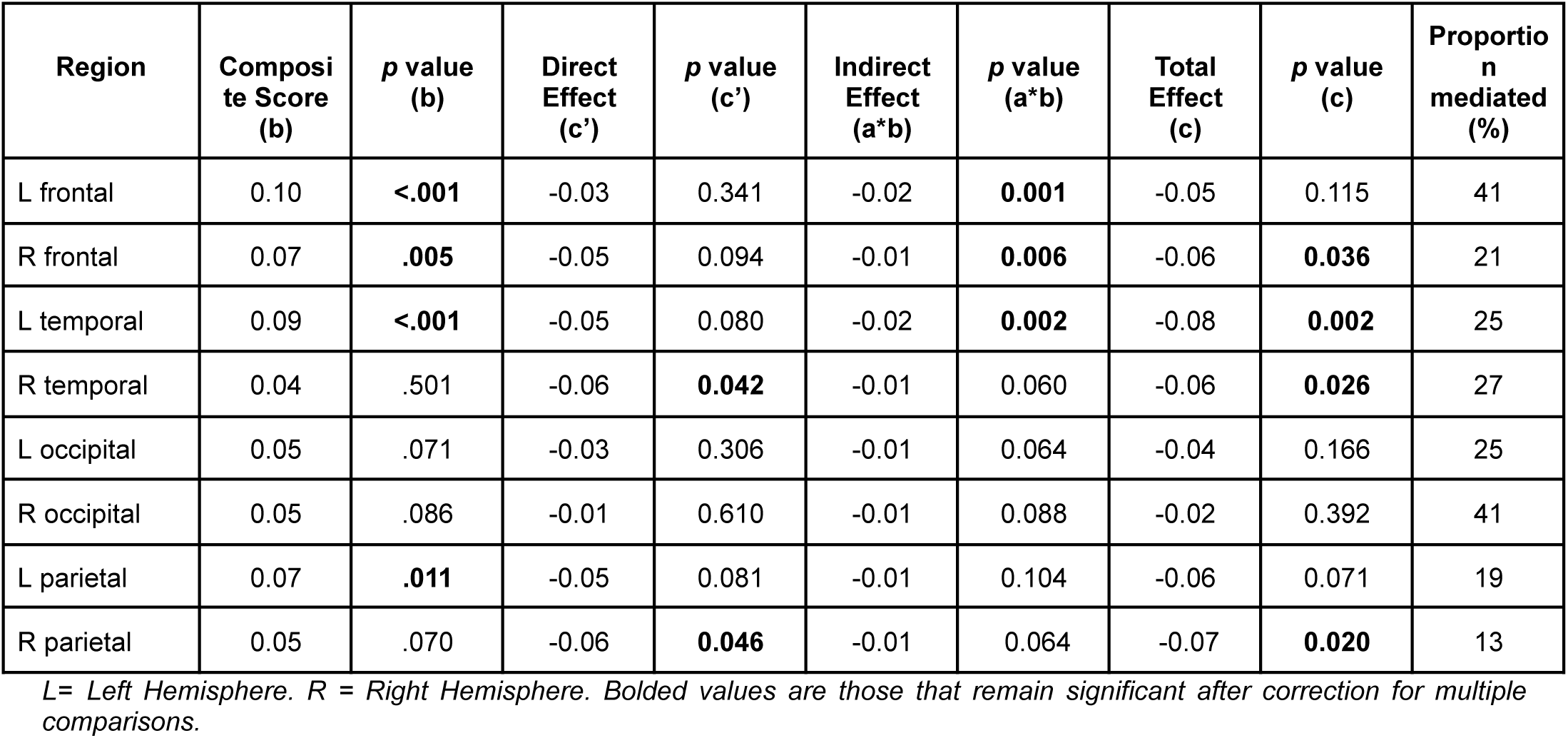
Mediation Analysis (Direct and Indirect Effects).

## 4 Discussion

This study investigated whether vascular risk factors mediate the relationship between educational attainment and WMH burden in older adults. Our findings demonstrate that higher education is significantly associated with reduced WMH burden across total and regional brain areas, with the most robust associations observed in the frontal, temporal, and parietal lobes. These findings align with the cognitive reserve hypothesis, supporting the idea that individuals with higher education levels may be better protected against the accumulation of cerebrovascular pathology (24,25). Cognitive reserve is theorized to enable individuals to maintain cognitive function despite brain pathology, potentially through more efficient or flexible brain networks or greater synaptic density. Higher education has consistently been associated with lower dementia risk, slower cognitive decline, and improved resilience to structural brain changes (26,58). The observed mediation by vascular risk factors suggests that part of the protective effect of education may occur through healthier cardiovascular profiles (59,60). These findings reinforce the notion that vascular risk partially mediates the relationship between education and WMH burden (9,10,24,58,60).

Our results showed that higher education is associated with lower levels of individual vascular risk factors, including systolic blood pressure, BMI, smoking, diabetes, hypertension, and hypercholesterolemia. This finding is consistent with population-level studies indicating that individuals with higher education are more likely to engage in health-promoting behaviors and experience better cardiovascular outcomes (26,26,30,59). Such behavioral and systemic advantages may contribute to reduced cerebrovascular burden and slower progression of WMHs (9,10,58). In our sample, education was not significantly associated with diastolic blood pressure or alcohol abuse, likely reflecting measurement limitations in the alcohol variable and a more stable profile of diastolic pressure in late life.

Second, education was inversely associated with WMH burden after adjusting for covariates, including diagnosis. This finding is in line with prior studies demonstrating that education contributes to cerebrovascular integrity and preserves white matter microstructure (24,26,61,62). Notably, the associations between education and WMHs were significant in anterior (frontal and temporal) and parietal regions but not occipital lobes, suggesting that the protective influence of education may be more pronounced in brain regions vulnerable to age- and vascular-related injury. This pattern aligns with Biesbroek et al. (53), who found that vascular risk factors were more strongly associated with anterior WMHs, while amyloid pathology explained variance in posterior regions. Similarly, Li et al. (58) reported that higher WMH burden was more strongly linked to neurodegeneration in posterior lobes, and that education moderated this relationship. These comparisons suggest that education may exert its greatest protective effects in vascularly vulnerable regions of the brain, particularly through attenuation of anterior WMH accumulation (22,63,64).

Third, mediation analysis revealed that vascular risk factors partially mediated the relationship between education and WMH burden, accounting for approximately 22% of the total effect. This finding is also consistent with previous studies that highlight the dual impact of education on brain health, directly through cognitive reserve and indirectly by promoting healthier vascular profiles (24,25). Regionally, mediation effects were most prominent in the frontal and temporal lobes. In the left and right frontal as well as the left temporal regions, only the indirect effects via vascular risk were significant, while direct effects were not, indicating a full mediation effect. No mediation effects were observed in the occipital and parietal lobes. This distribution aligns with studies showing that anterior WMHs are more susceptible to vascular risk (53,65,66), while posterior WMHs are typically associated with neurodegenerative pathology, particularly amyloid and tau accumulation (21,60,67). This finding suggests that vascular risk factors exert region-specific effects on WMH accumulation, and that the benefits of higher education may be most evident in brain areas more susceptible to modifiable vascular damage (20,21,53,58). This topographical differentiation reinforces the need for region-specific WMH analyses, as anterior regions, unlike posterior regions which may reflect more static, neurodegeneration-related damage, appear more responsive to vascular intervention. Collectively, these comparisons suggest that lobar-level mediation analysis provides additional insights into how vascular and neurodegenerative processes differentially shape WMH burden (9,10,21,53,59,60). The presence of partial mediation in anterior lobes underscores the potential for targeted vascular health interventions to mitigate WMH accumulation, particularly among individuals with lower educational attainment.

Several limitations in the current study warrant consideration. The cross-sectional design precludes causal inference, and prospective studies are needed to evaluate the longitudinal mediation effects of vascular risk on education-related differences in WMH accumulation. Additionally, our main analyses were limited to White participants due to insufficient representation in other racial and ethnic groups. While we reported the findings for other racial and ethnic groups, these results were likely underpowered to detect significant associations in these populations, and future studies with sufficiently powered and more diverse samples are essential to enhance generalizability. Participants in the NACC are volunteers who are often more educated and healthier than the general population, which could further limit the generalizability of our findings. We also note limitations in our assessment of alcohol abuse. We relied solely on the ALCOHOL variable, which defines alcohol abuse as causing clinically significant impairment in areas such as work, driving, legal, or social domains over a 12-month period. Other variables related to alcohol intake contained excessive missing data and could not be reliably included in our models. Moreover, some of the vascular risk factors were modeled as binary indicators, which may not fully capture the complexity of vascular health; future work should incorporate more detailed clinical and laboratory-based measures of vascular function. Lastly, education was used as a proxy for cognitive reserve but may not capture broader life experiences that contribute to neural resilience.

In conclusion, our study supports the hypothesis that education influences WMH burden by reducing vascular disease burden. The findings underscore the need for public health strategies that promote both educational access and vascular health management as potential levers to reduce cerebrovascular disease and cognitive aging risk.

## Data Availability

The data utilized in this study were also sourced from the National Alzheimer's Coordinating Center (NACC) database (https://naccdata.org/), specifically drawing from the NACC Uniform Data Set (UDS) and MRI Data Set (Beekly et al., 2004; Besser, Kukull, Knopman, et al., 2018; Besser, Kukull, Teylan, et al., 2018).

## Acknowledgments

The NACC database is funded by NIA/NIH Grant U24 AG072122. NACC data are contributed by the NIAfunded ADRCs: P30 AG062429 (PI James Brewer, MD, PhD), P30 AG066468 (PI Oscar Lopez, MD), P30 AG062421 (PI Bradley Hyman, MD, PhD), P30 AG066509 (PI Thomas Grabowski, MD), P30 AG066514 (PI Mary Sano, PhD), P30 AG066530 (PI Helena Chui, MD), P30 AG066507 (PI Marilyn Albert, PhD), P30 AG066444 (PI David Holtzman, MD), P30 AG066518 (PI Lisa Silbert, MD, MCR), P30 AG066512 (PI Thomas Wisniewski, MD), P30 AG066462 (PI Scott Small, MD), P30 AG072979 (PI David Wolk, MD), P30 AG072972 (PI Charles DeCarli, MD), P30 AG072976 (PI Andrew Saykin, PsyD), P30 AG072975 (PI Julie A. Schneider, MD, MS), P30 AG072978 (PI Ann McKee, MD), P30 AG072977 (PI Robert Vassar, PhD), P30 AG066519 (PI Frank LaFerla, PhD), P30 AG062677 (PI Ronald Petersen, MD, PhD), P30 AG079280 (PI Jessica Langbaum, PhD), P30 AG062422 (PI Gil Rabinovici, MD), P30 AG066511 (PI Allan Levey, MD, PhD), P30 AG072946 (PI Linda Van Eldik, PhD), P30 AG062715 (PI Sanjay Asthana, MD, FRCP), P30 AG072973 (PI Russell Swerdlow, MD), P30 AG066506 (PI Glenn Smith, PhD, ABPP), P30 AG066508 (PI Stephen Strittmatter, MD, PhD), P30 AG066515 (PI Victor Henderson, MD, MS), P30 AG072947 (PI Suzanne Craft, PhD), P30 AG072931 (PI Henry Paulson, MD, PhD), P30 AG066546 (PI Sudha Seshadri, MD), P30 AG086401 (PI Erik Roberson, MD, PhD), P30 AG086404 (PI Gary Rosenberg, MD), P20 AG068082 (PI Angela Jefferson, PhD), P30 AG072958 (PI Heather Whitson, MD), P30 AG072959 (PI James Leverenz, MD).

## Consent Statement

Informed consent was obtained in writing from each participant or their designated study partner.

## Contributions

S.R, R.M, Y.Z, M.D, and C.M, were involved with the conceptualization and design of the work. S.R, Y.Z and M.D. completed analysis and C.M, M.D, R.M, Y.Z, and S.R were involved with data interpretation. S.R wrote the manuscript. and C.M, R.M, Y.Z, M.D, and S.R revised and approved the submitted version.

## Supplementary Materials

**Table S1:** Demographic, Clinical, and Neuroimaging Characteristics of Black Participants.

|  | Total | NC | MCI | AD |
| --- | --- | --- | --- | --- |
| Total number | 143 | 61 | 60 | 22 |
| Age (Mean, SD) | 76.04 (8.88) | 73.55 (8.60) | 77.61 (8.55) | 78.67 (9.19) |
| Female (n, %) | 99 (69) | 48 (79) | 36 (60) | 15 (68) |
| Education (Years, Mean, SD) | 14.32 (3.38) | 14.61 (3.25) | 14.58 (3.32) | 12.82 (3.66) |
| Diabetes (n, %) | 54 (38) | 23 (38) | 28 (47) | 3 (14) |
| Hypertension (n, %) | 101 (71) | 42 (69) | 47 (78) | 12 (54) |
| Hypercholesterolemia (n, %) | 84 (59) | 32 (52) | 40 (67) | 12 (54) |
| Alcohol abuse (n, %) | 5 (3) | 0 | 4 (6) | 1 (4) |
| Smoking (n, %) | 63 (44) | 26 (43) | 28 (47) | 9 (41) |
| High BMI ($\geq 25$ kg/m<sup>2</sup>) (n, %) | 101 (71) | 48 (79) | 37 (62) | 16 (73) |
| High systolic BP ($\geq 130$ mm Hg) (n, %) | 103 (72) | 42 (69) | 46 (77) | 15 (68) |
| High diastolic BP ($\geq 80$ mm Hg) (n, %) | 59 (41) | 26 (43) | 25 (42) | 8 (36) |
| Total WMH Volume FLAIR (mm<sup>3</sup>) (Mean, SD) | 27234.32<br>(22987.10) | 19985<br>(19935) | 33138<br>(24214) | 31233<br>(22833) |
| Composite vascular score (Mean, D) | 3.40 (1.55) | 3.42 (1.48) | 3.59 (1.65) | 2.96 (1.69) |

NC = Cognitively Normal. MCI = Mild Cognitive Impairment. AD = Alzheimer's disease. SD = Standard Deviation. BP = Blood Pressure. BMI = Body Mass Index. WMH = White matter hyperintensity. Values are presented as total number and percentage or mean and standard deviation. Diabetes, hypertension, hypercholesterolemia, alcohol abuse, and smoking were reported as 1 (has the condition) or 0 (does not have the condition). BMI, systolic and diastolic blood pressure were continuous measures.

**Table S2:** Demographic, Clinical, and Neuroimaging Characteristics of Hispanic Participants.

|  | Total | NC | MCI | AD |
| --- | --- | --- | --- | --- |
| Total number | 171 | 67 | 52 | 62 |
| Age (Mean, SD) | 74.87 (8.47) | 72.61 (7.14) | 76.32 (9.27) | 76.33 (8.74) |
| Female (n, %) | 108 (63) | 43 (64) | 30 (58) | 35 (56) |
| Education (Years, Mean, SD) | 10.64 (4.93) | 11.70 (4.44) | 10.40 (5.26) | 9.52 (5.01) |
| Diabetes (n, %) | 54 (31) | 22 (33) | 18 (35) | 14 (22) |
| Hypertension (n, %) | 114 (67) | 44 (66) | 37 (71) | 33 (53) |
| Hypercholesterolemia (n, %) | 96 (56) | 37 (55) | 31 (60) | 28 (45) |
| Alcohol abuse (n, %) | 15 (9) | 5 (7) | 5 (9) | 5 (8) |
| Smoking (n, %) | 74 (43) | 29 (43) | 27 (52) | 18 (29) |
| High BMI ($\geq 25$ kg/m<sup>2</sup>) (n, %) | 127 (74) | 54 (80) | 37 (71) | 36 (58) |
| High systolic BP ($\geq 130$ mm Hg) (n, %) | 119 (69) | 47 (70) | 38 (73) | 34 (55) |
| High diastolic BP ($\geq 80$ mm Hg) (n, %) | 63 (37) | 24 (36) | 15 (29) | 24 (39) |
| Total WMH Volume FLAIR (mm<sup>3</sup>) (Mean, SD) | 19817.82 (17963.50) | 14951 (16836) | 22635 (18361) | 23271 (17897) |
| Composite vascular score (Mean, D) | 3.29 (1.55) | 3.35 (1.51) | 3.29 (1.49) | 3.21 (1.69) |

NC = Cognitively Normal. MCI = Mild Cognitive Impairment. AD = Alzheimer's disease. SD = Standard Deviation. BP = Blood Pressure. BMI = Body Mass Index. WMH = White matter hyperintensity. Values are presented as total number and percentage or mean and standard deviation. Diabetes, hypertension, hypercholesterolemia, alcohol abuse, and smoking were reported as 1 (has the condition) or 0 (does not have the condition). BMI, systolic and diastolic blood pressure were continuous measures.

**Table S3:** Demographic, Clinical, and Neuroimaging Characteristics of Asian Participants.

|  | Total | NC | MCI | AD |
| --- | --- | --- | --- | --- |
| Total number | 40 | 13 | 21 | 6 |
| Age (Mean, SD) | 77.40 (8.77) | 75.25 (7.74) | 78.40 (9.60) | 78.60 (8.33) |
| Female (n, %) | 25 (62) | 6 (46) | 14 (67) | 5 (83) |
| Education (Years, Mean, SD) | 15.25 (2.99) | 16 (2.83) | 15.33 (3.24) | 13.33 (1.50) |
| Diabetes (n, %) | 12 (30) | 5 (38) | 7 (33) | 0 |
| Hypertension (n, %) | 23 (57) | 7 (54) | 12 (57) | 4 (67) |
| Hypercholesterolemia (n, %) | 24 (60) | 7 (54) | 15 (71) | 2 (33) |
| Alcohol abuse (n, %) | 1 (2) | 0 | 1 (5) | 0 |
| Smoking (n, %) | 9 (22) | 4 (31) | 2 (9) | 3 (50) |
| High BMI ($\geq 25$ kg/m<sup>2</sup>) (n, %) | 15 (37) | 4 (31) | 9 (43) | 2 (33) |
| High systolic BP ($\geq 130$ mm Hg) (n, %) | 25 (62) | 5 (38) | 16 (76) | 4 (67) |
| High diastolic BP ($\geq 80$ mm Hg) (n, %) | 11 (27) | 3 (23) | 4 (19) | 4 (67) |
| Total WMH Volume FLAIR (mm<sup>3</sup>) (Mean, SD) | 24945.75 (16661.62) | 18114 (16206) | 27341 (16571) | 31363 (15548) |
| Composite vascular score (Mean, D) | 2.75 (1.60) | 2.38 (1.76) | 3 (1.45) | 2.67 (1.86) |

NC = Cognitively Normal. MCI = Mild Cognitive Impairment. AD = Alzheimer's disease. SD = Standard Deviation. BP = Blood Pressure. BMI = Body Mass Index. WMH = White matter hyperintensity. Values are presented as total number and percentage or mean and standard deviation. Diabetes, hypertension, hypercholesterolemia, alcohol abuse, and smoking were reported as 1 (has the condition) or 0 (does not have the condition). BMI, systolic and diastolic blood pressure were continuous measures.

**Table S4:** Associations Between Educational Attainment and Individual Vascular Risk Factors in Non-White Participants.

|  | Black |  |  | Hispanic |  |  | Asian |  |  |
| --- | --- | --- | --- | --- | --- | --- | --- | --- | --- |
|  | Estimate | tStat | *p* value | Estimate | tStat | *p* value | Estimate | tStat | *p* value |
| Diabetes | -0.01 | -0.18 | 0.85 | -0.05 | -2.59 | **0.01** | -0.12 | -1.03 | 0.31 |
| Hypertension | -0.02 | -0.41 | 0.68 | -0.06 | -2.79 | **0.01** | -0.05 | -0.56 | 0.58 |
| Systolic Blood Pressure | 0.00 | -0.10 | 0.92 | -0.03 | -1.97 | 0.05 | -0.07 | -1.32 | 0.20 |
| Diastolic Blood Pressure | 0.01 | 0.44 | 0.66 | 0.00 | 0.15 | 0.88 | -0.04 | -0.83 | 0.41 |
| Hypercholesterolemia | -0.02 | -0.55 | 0.58 | -0.43 | -4.72 | **<0.001** | 0.00 | 0.04 | 0.97 |
| Alcohol Abuse | -0.02 | -0.60 | 0.55 | -0.04 | -3.07 | **<0.001** | -0.04 | -0.84 | 0.41 |
| Smoking (Years) | -0.05 | -1.89 | 0.06 | -0.04 | -2.34 | **0.02** | 0.03 | 0.76 | 0.45 |
| Smoking (Packs Per Day) | -0.04 | -1.79 | 0.08 | -0.01 | -0.97 | 0.33 | -0.02 | -0.41 | 0.69 |
| BMI | -0.02 | -0.90 | 0.37 | -0.05 | -3.14 | **<0.001** | -0.05 | -1.19 | 0.24 |
| Composite Score | -0.05 | 0.53 | 0.60 | -0.11 | -1.65 | 0.10 | -0.01 | -3.94 | 0.08 |

BMI = Body Mass Index. Bolded values are those that remained significant after correction for multiple comparisons.

**Table S5:**
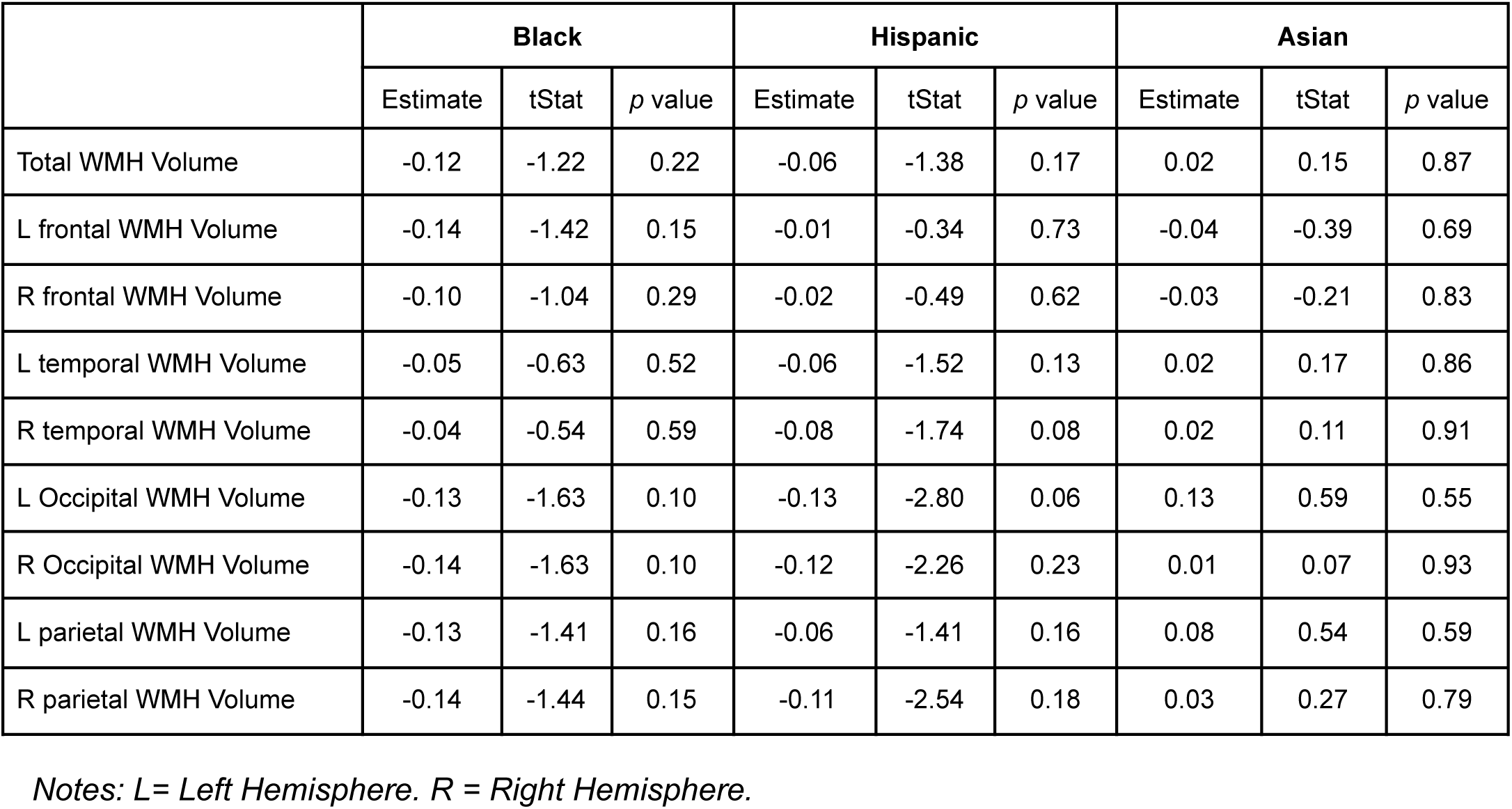
Associations Between Education and Total and Regional WMH Volumes in Non-White Participants.

|  | Black |  |  | Hispanic |  |  | Asian |  |  |
| --- | --- | --- | --- | --- | --- | --- | --- | --- | --- |
|  | Estimate | tStat | *p* value | Estimate | tStat | *p* value | Estimate | tStat | *p* value |
| Total WMH Volume | -0.12 | -1.22 | 0.22 | -0.06 | -1.38 | 0.17 | 0.02 | 0.15 | 0.87 |
| L frontal WMH Volume | -0.14 | -1.42 | 0.15 | -0.01 | -0.34 | 0.73 | -0.04 | -0.39 | 0.69 |
| R frontal WMH Volume | -0.10 | -1.04 | 0.29 | -0.02 | -0.49 | 0.62 | -0.03 | -0.21 | 0.83 |
| L temporal WMH Volume | -0.05 | -0.63 | 0.52 | -0.06 | -1.52 | 0.13 | 0.02 | 0.17 | 0.86 |
| R temporal WMH Volume | -0.04 | -0.54 | 0.59 | -0.08 | -1.74 | 0.08 | 0.02 | 0.11 | 0.91 |
| L Occipital WMH Volume | -0.13 | -1.63 | 0.10 | -0.13 | -2.80 | 0.06 | 0.13 | 0.59 | 0.55 |
| R Occipital WMH Volume | -0.14 | -1.63 | 0.10 | -0.12 | -2.26 | 0.23 | 0.01 | 0.07 | 0.93 |
| L parietal WMH Volume | -0.13 | -1.41 | 0.16 | -0.06 | -1.41 | 0.16 | 0.08 | 0.54 | 0.59 |
| R parietal WMH Volume | -0.14 | -1.44 | 0.15 | -0.11 | -2.54 | 0.18 | 0.03 | 0.27 | 0.79 |

*Notes: L= Left Hemisphere. R = Right Hemisphere.*

